# ‘Look up from the waiting list and see the bigger picture’: A qualitative analysis of clinical specialist physiotherapist perspectives on low back pain care in Ireland

**DOI:** 10.1101/2023.11.15.23298561

**Authors:** Cathriona Murphy, Helen French, Geraldine McCarthy, Caitriona Cunningham

## Abstract

**Introduction:** Healthcare systems are struggling to deliver high-quality low back pain (LBP) care. In 2012 specialist physiotherapist-led musculoskeletal (MSK) triage services were introduced in Irish hospitals to expedite patient care and alleviate pressure on elective orthopaedic/ rheumatology consultant clinics. Specialist physiotherapists have expertise to inform health service improvement and reform, but their perspectives of LBP healthcare delivery have received scant attention.

**Objectives:** To explore specialist physiotherapists’ perspectives on LBP care in Ireland, the barriers and facilitators to quality LBP care and the development of MSK interface services in primary care settings.

**Design:** Cross-sectional observational study using an anonymous electronic survey with thematic framework analysis of response data from open-ended questions.

**Participants:** Thirty-four clinical specialist physiotherapists in Irish MSK triage services.

**Results:** Thematic analysis resulted in six overarching themes, grouped into two categories. One category pertained to LBP healthcare in Ireland with the following three themes: 1) Inadequate health services for patients with LBP; 2) Need for defined LBP clinical pathways; 3) Need for a multisectoral approach to spine health. Themes in the second category, pertaining to the development of community-based MSK interface services, were: 4) Concern regarding isolation from secondary care services; 5) Unrealistic expectations of MSK triage; 6) Improved communication and collaboration with primary care services.

**Conclusion:** Specialist physiotherapists have concerns regarding LBP health services and persistence of a biomedical, secondary care-led approach. They advocate for investment in primary care multi-disciplinary teams, enhanced integration across primary and secondary care, development of a national clinical pathway and a multisectoral approach.

## Introduction

Low back pain (LBP), the primary cause of years lived with disability [1, 2], remains a substantial global health problem, placing a considerable burden on individuals, healthcare systems and the wider socioeconomic milieu. Despite largely consistent best practice clinical guidelines, an evidence-practice gap remains [3], indicating that healthcare systems are struggling to deliver high-quality LBP care whilst caring for rising numbers presenting with MSK disorders due to a growing and ageing global population [4].

In Ireland the increased burden of MSK disorders has contributed to long wait times for orthopaedic and rheumatology speciality care, further compounded by insufficient consultant resources. Ireland has 2.6 orthopaedic consultants per 100,000 population [5] compared to 8 in the UK [6] and one of the lowest ratios of rheumatologists to population in the European Union [7]. For the person with LBP, General Practitioners (GPs) are usually the first contact point and gatekeepers to elective Irish public healthcare services, with onward referral options including primary care (PC) physiotherapy and secondary care (SC) consultant-led orthopaedic and rheumatology services. In 2011 the Health Service Executive (HSE), the organisation responsible for Irish public health service provision, developed the National MSK Physiotherapy Triage Initiative to address long outpatient waiting lists for orthopaedic/ rheumatology consultant services. This initiative sees patients on these waiting lists being assessed by clinical specialist (CS) physiotherapists, experienced practitioners with advanced skills in management of MSK conditions, in hospital-based MSK triage clinics under clinical governance of orthopaedic/ rheumatology consultants. Similar to advanced practice physiotherapist triage services in other countries, these triage services manage patient care without onward referral for consultant review in > 80% of cases [8, 9]. The high proportion discharged from Irish MSK triage clinics suggests that many patients are unnecessarily referred to SC consultant services, contributing to unnecessarily long wait times. This is contrary to LBP best practice guidelines, which consistently advocate a demedicalised approach in PC settings for most patients [3, 10]. In line with the HSE’s objective to optimise community-based care [11], the National MSK Initiative has recently increased CS physiotherapist MSK triage posts from 30 to 65 to facilitate expansion to community-based interface clinics.

Utilising clinicians’ experience and knowledge to inform the planning, delivery and implementation of health service reform is critical for successful quality improvement initiatives, but meaningful clinician engagement is a weakness in health service development in Ireland and more broadly [12–15]. Health and social care professionals (HSCPs), also known as allied health professionals, are underrepresented in health leadership roles, further limiting their ability to instigate or influence organisational change and healthcare reform [16]. Specialist MSK physiotherapists are subject matter experts in LBP, but there has been little documented on their views of health system LBP care and none in the Irish context. Marking 10 years of MSK triage services (2012-2022) and to inform the development of Irish LBP services, we conducted a national survey of MSK triage CS physiotherapists with the following aims: 1) to explore their perceptions of LBP care in Ireland, including barriers and facilitators to quality care; 2) to establish how MSK triage services function with respect to the delivery of LBP care; 3) to examine opinions on the development of community-based MSK interface clinics. The survey included both closed- and open-ended questions. Here we present a thematic qualitative analysis of data from seven open-ended questions (supplementary material 1).

## Methods

### Study Design

This study employed a cross-sectional observational design using a bespoke, anonymous electronic survey. Qualitative analysis is reported in accordance with Standards for Reporting Qualitative Research (SRQR) [17].

### Survey design and distribution

Survey development included piloting amongst four CS physiotherapists. Survey questions were constructed on Qualtrics XM (Qualtrics XM, Utah, US) and grouped in five sections, with open questions eliciting opinions on (i) LBP care delivery in Ireland (ii) MSK triage service links with PC services and (iii) development of community-based MSK LBP triage services. The survey was distributed to all National MSK Initiative CS physiotherapists via the Initiative’s email distribution list, along with participant information regarding the research and its purpose. Informed consent was obtained. The survey remained active for 11 weeks; one reminder email was sent.

### Data management and analysis

Survey data were exported from Qualtrics XM into Microsoft Excel for descriptive statistical analysis. Free text responses were imported from Microsoft Excel to NVivo 12 software (QSR International Pty Ltd.) for qualitative data analysis.

Qualitative data analysis was guided by framework analysis [18]. Analysis was informed by critical realist ontology and contextualist epistemology, reflecting the researchers’ understanding that the knowledge generated is contextual with an interpretative element, but reflects an underlying truth [19]. Following familiarisation with the free text responses, the data of 10 participants were independently and inductively coded line-by-line in NVivo by two research team members (CM; CC). CM is an MSK triage CS physiotherapist and PhD candidate; CC is an Associate Professor of Physiotherapy and an experienced researcher. Preliminary codes were discussed by CM and CC to resolve discrepancies and produce an agreed analytic framework of codes that was subsequently applied to the remaining dataset by CM. This line-by-line indexing of the remaining dataset to the analytic framework allowed for the addition of new codes, but none were required. CM collated all indexed data into a framework matrix before collating similar preliminary subthemes into groups to develop preliminary themes. Preliminary themes and subthemes were then refined (CM; CC) in an iterative process to develop overarching themes with a higher level of abstraction. These themes were further refined and agreed by whole group discussion.

### Validity and reflexivity

Open coding by two independent research team members contributed to validity. Theme refinement was conducted with continual reference to the data to ensure that overarching themes remained reflective of participants’ opinions. The lead author, conscious of potential bias from her clinical role, completed a reflective exercise prior to data analysis through answering the open questions and revisiting these answers following final theme development to aid critical reflection and enhance trustworthiness of the research process.

## Results

The eligible sample, excluding CS physiotherapists on long-term leave and those that don’t accept referrals for LBP, was n=53. Thirty-four physiotherapists answered the open questions (response rate 64%). Most respondents were female (82.4%; n=28) and 56% reported > 20 years’ experience (n=19; range 6 to > 20 years).

### Themes

Data analysis resulted in six overarching themes, which were grouped into two categories: A) LBP healthcare in Ireland (themes 1-3) and B) development of community-based MSK interface services (themes 4-6). Tables 1 and 2 contain illustrative quotes for these themes.

**Table 1:**
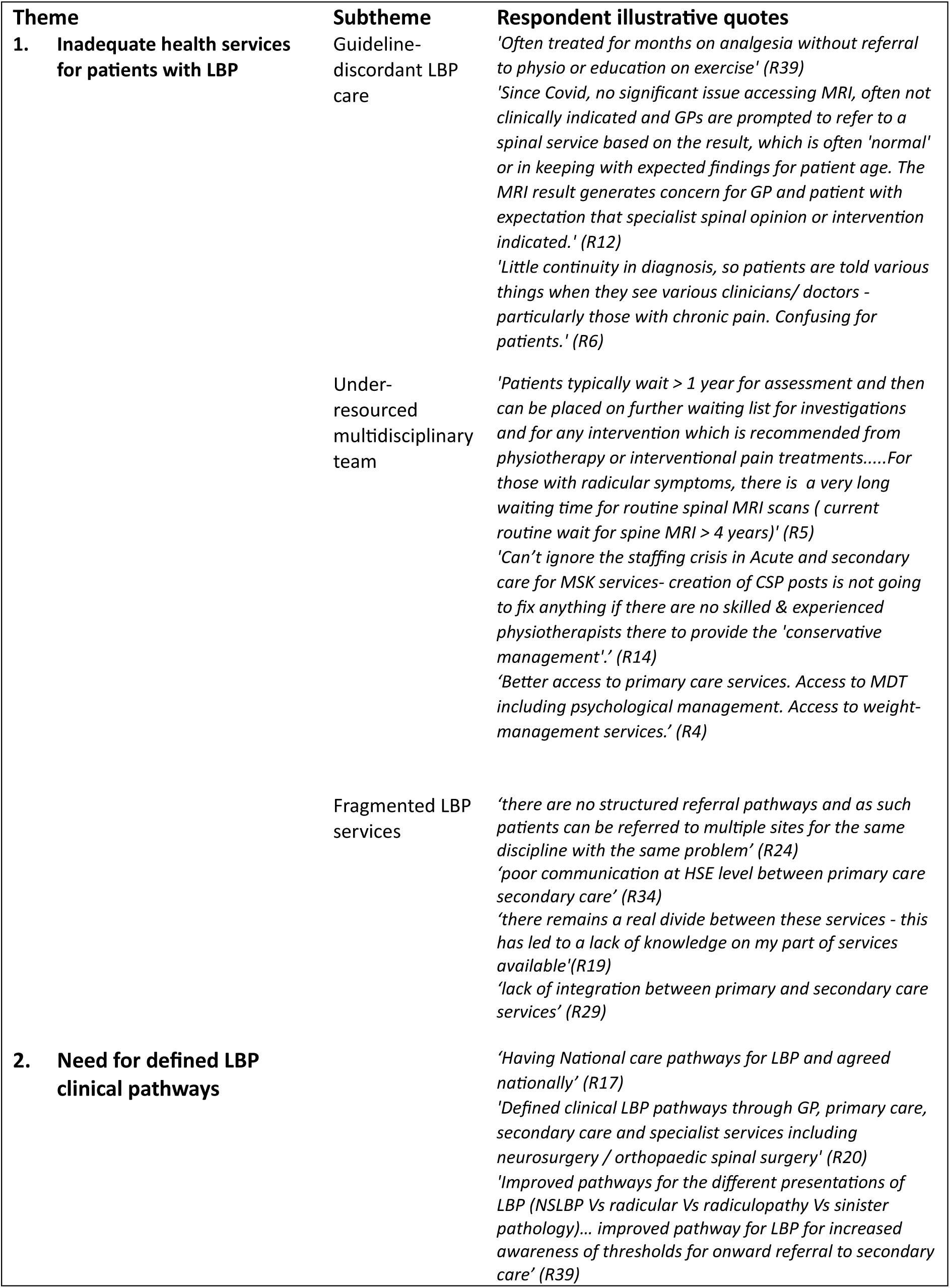
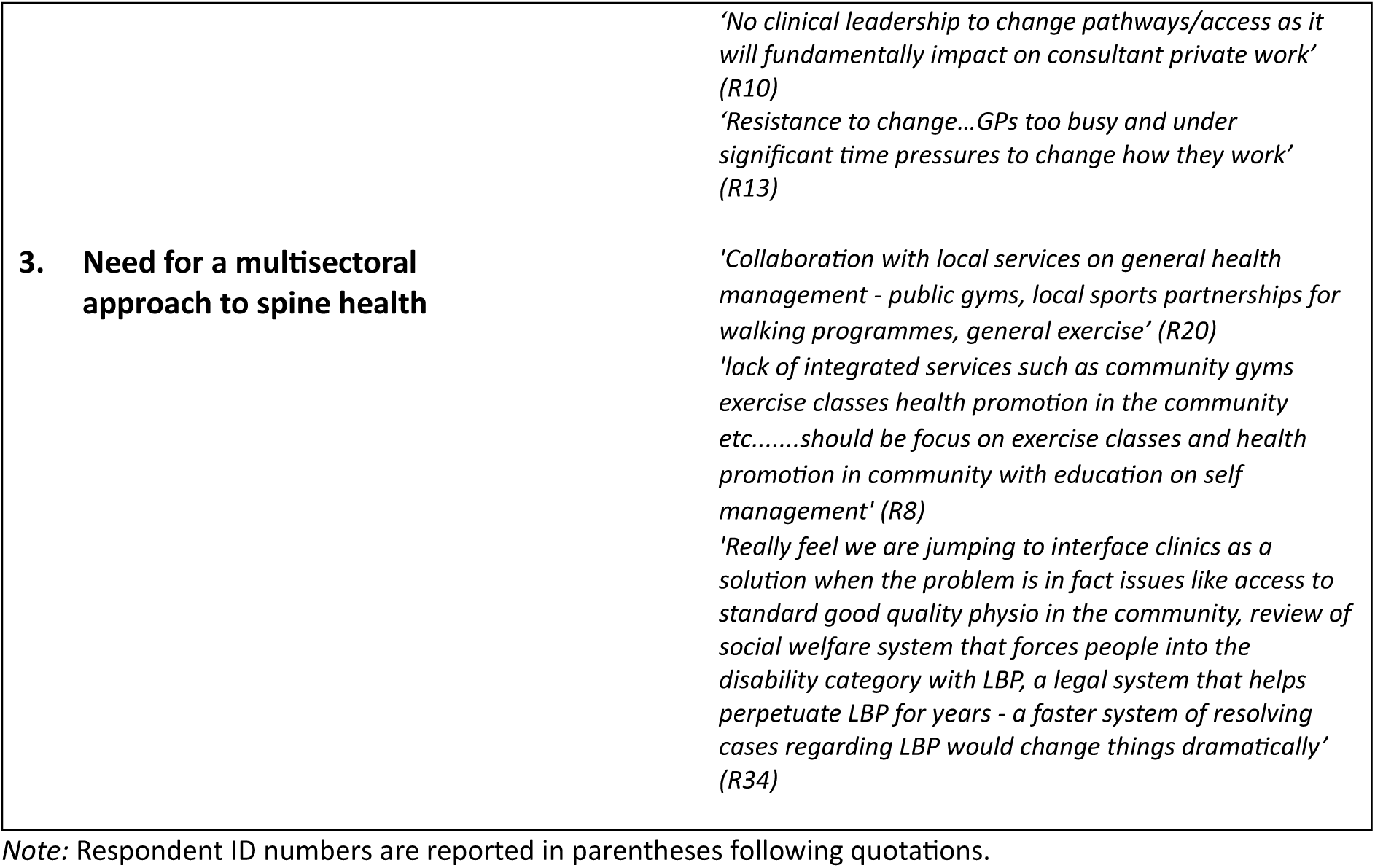
Quotes illustrating themes related to clinical specialist physiotherapists’ views on LBP healthcare in Ireland.

**Table 2:** Quotes illustrating themes related to clinical specialist physiotherapists’ views on the development of community-based MSK interface services in Ireland.

| Theme | Respondent illustrative quotes |
| --- | --- |
| 4. **Concern regarding isolation from secondary care services** | *'No physiotherapist should work solely in interface. They should also always maintain clinics working alongside the consultant. We need to maintain formal communication channels with consultants and the medical team. There is a danger that these will be lost due to other demands on consultants' time....after a number of years working alongside orthopaedic teams, MSKs are a great resource - we are in great danger of obliterating this by forcing physios to work on their own in a room trudging through a never-ending waiting list. Use your MSKs better, look up from the waiting list and see the bigger picture.'* (R34)<br>*'risk of been 'forgotten' by the consultant when off site, lack of support and difficulty discussing cases with consultant and accessing second opinion'* (R27)<br>*'Dilution of the excellent team based approach to care within an embedded triage service in the consultants' clinics'* (R31) |
| 5. **Unrealistic expectations of community MSK Triage clinics** | *'We have to cover all specialities - upper limb clinics lower limb clinics and within these clinics see our LBP patients. we are expected to be masters of everything and provide the support and follow up for these patient without returning them to our clinic. This is with the lack an integrated pathway of care in community with excessive waiting times for follow up and huge resource pressures in the community..... The reality is community based MSK interface clinics are physio only and we cannot provide everything'* (R32)<br>*'Current CSP triage physiotherapist to specialist in LBP- rather than having to triage all joints in ortho and Rheum. Too difficult to specialise in every joint and develop integrated pathways between acute and primary care for specific conditions at the same time.'* (R14)<br>*'We do not enjoy the infrastructure and the team structure at interface that the NHS has. We also do not have formal structures re education and development of staff. We need to think again before we send sole practitioner physiotherapists out to a room in a remote primary care centre and expect them to be the panacea of all ills.'* (R34) |
| 6. **Improved communication and collaboration with primary care services** | *'Better communication and learning between the services - for secondary care to be aware of different services available, easier referral process. For primary care better understanding of spinal pathways, need for investigation etc'* (R19)<br>*'Improved e-referral/open links of communication. Improved discharge/update summaries. Improved communication regarding primary care wait times, waiting list criteria and understanding of treatment pathways'* (R24)<br>*'during current expansion to interface/outreach clinics as per NCP and recent local MSK personnel expansion, we are delivering quarterly inservice training to primary care physiotherapists, and have established an open line of communication between MSK in secondary care and physiotherapy in primary care'* (R20)<br>*'Currently working on building relationships with primary care teams and integrating clinics onsite'* (R37) |

Note: Respondent ID numbers are reported in parentheses following quotations.

### LBP healthcare in Ireland

#### 1. Inadequate health services for patients with LBP

Respondents expressed concerns that patients with LBP frequently experience inadequate services, characterised by the following subthemes: guideline-discordant care; fragmented services; and an under-resourced multi-disciplinary team (MDT).

Respondents highlighted guideline-discordant care with a biomedical orientation, including over-reliance on imaging and medication, unnecessary SC referral without adequate first-line treatments, and under-emphasis on exercise rehabilitation, education and self-management. Inconsistent information from healthcare professionals was identified as generating confusion for patients.

Physiotherapists considered the MDT for LBP care to be under-resourced, particularly PC services, which were perceived as overwhelmed and of low priority for the HSE. Limited or no access to weight management, psychological, occupational therapy and pain management services were reported, as well as long wait times for MSK triage, Magnetic Resonance Imaging (MRI), physiotherapy, consultant appointments, injections and pain management services. Long wait times were identified as negatively impacting patients’ physical and emotional wellbeing, as well as respondents’ perception of the effectiveness of their role, citing lack of value in moving patients from one waiting list to another.

Respondents identified current LBP service provision as fragmented, inefficient and lacking structure, and described poor communication and integration between PC and SC at clinician and organisational levels, leading to a knowledge gap for clinicians regarding available services.

#### 2. Need for defined LBP clinical pathways

Defined clinical pathways across PC, SC and specialist services were seen as a means of service improvement. Suggestions for inclusion in LBP clinical pathways included: defined referral guidelines; returning inappropriate referrals to referrers; care stratification based on risk of chronicity; direct referral from GPs and PC physiotherapists to MSK triage; emphasis on PC services to reduce unnecessary SC referral; national implementation of pathways for consistency and compliance. Clinical pathways should also address the perceived disadvantage (delayed care with more limited management option) to patients accessing triage at clinical sites without spine surgical services, when compared to those accessing triage with co-located surgical services. Despite advocating for clinical pathways, respondents expressed reservations about the health service’s capacity for change, citing organisational resistance, lack of incentive, poor cooperation between PC and SC, potential conflict of interest between private and public healthcare, and lack of clinical leadership.

#### 3. Need for a multisectoral approach to spine health

Respondents communicated the need for a multisectoral approach to spine health to optimise outcomes. Community exercise promotion through collaboration with public gyms and community groups was emphasised, as well as the need to address social welfare and legal system influences, which were seen as contributing to LBP-associated disability.

### Development of community-based MSK interface services

#### 4. Concern regarding isolation from SC services

Respondents expressed concern that moving MSK triage to community-based services would result in isolation from SC services with adverse consequences for clinical and operational efficiency through impaired team working, reduced consultant support, delayed decision making and reduced opportunities for on-the-job learning, particularly for new appointees. Respondents valued colocation with consultant teams for facilitating communication, team working and learning, with team members having different but complementary skills. Lack of same-day consultant opinion and radiology services in PC was seen as potentially leading to excessive review appointments in SC. Consultant support of community-based triage was regarded as essential for successful implementation, but some respondents felt this was lacking. Appropriate IT infrastructure and shared electronic records between PC and SC were considered necessary to manage logistical issues regarding access to medical charts and record keeping. Respondents emphasised the need for triage physiotherapists to avoid working solely in PC settings.

#### 5. Unrealistic expectations of MSK triage services

Respondents conveyed being overburdened by unrealistic expectations of the clinical and operational service that can be provided by community-based MSK triage clinics. They perceive an expectation to have specialist knowledge across the plethora of MSK conditions, to efficiently manage patient care despite long wait times and lack of ancillary services, and to deliver operational change. Respondents felt unsupported in meeting these expectations, citing little formal, funded staff education and development, with training predominantly self-directed, self-funded and undertaken in their own time. Lone triage physiotherapists working in community settings cannot, in their opinion, facilitate substantial service change.

#### 6. Improved communication and collaboration with PC services

Respondents regarded enhanced collaboration with GPs and other PC providers as important to improve LBP healthcare and see the establishment of community-based triage services as an opportunity for improved communication, integration, and mutual understanding of available services across PC and SC.

## Discussion

Through collection and synthesis of CS physiotherapists’ perspectives on LBP healthcare in Ireland, this study enables better understanding of key issues and identifies improvement opportunities at micro (clinician), meso (health service/organisation) and macro (health and social care systems) health system levels. Key themes portray patients with LBP frequently receiving inadequate healthcare services characterised by treatment inconsistent with best practice, fragmented services and insufficiently staffed physiotherapy and wider multidisciplinary services. The analysis demonstrates that at micro level clinicians need to ensure consistent, evidence-informed care; at meso level improved care integration and communication is required across PC and SC supported by integrated IT systems; at the macro level national clinical pathways, funding of comprehensive PC MDTs, and a multisectoral approach should be facilitated.

Reports of guideline-discordant LBP care in Ireland are consistent with a persistent evidence-practice gap internationally [3], despite evidence that guideline-adherent care is more cost-effective and leads to improved outcomes [20, 21]. Barriers to clinical guideline adherence have been documented, and although solutions have been proposed, including defunding of ineffective investigations/ treatments and improved communication of the evidence base, context-specific implementation research is required to optimise knowledge translation to clinical practice [10, 22, 23].

Respondents portrayed a predominantly reactive, biomedical, SC-led model of LBP care that is not resourced to support demedicalised care in community settings. Their reports of insufficient physiotherapy resources are consistent with per capita physiotherapy supply in Ireland being 30% lower than EU-28 average [24]. It is concerning that patients with more complex needs were perceived to be particularly poorly served through inadequate access to skilled multidisciplinary care, as these patients, although small in number, account for the majority of LBP-associated disability and costs [25]. This under-development of multidisciplinary care is consistent with global under-recognition of the societal burden of LBP and MSK conditions at health policy level and an associated mismatch in allocated resources [26, 27].

The theme concerning the need for clinical pathways reflects respondents’ beliefs that such pathways implemented nationally may address service fragmentation and improve care coordination, quality, consistency and reduce geographic inequity. The Global Spine Care Initiative advocates for enhanced collaboration and integration in spine care [28] and end-to-end clinical pathways are proposed to support integration and best-practice implementation [3]. In a recent systematic review, LBP clinical pathways were associated with improved efficiency of care delivery, but their capacity to enhance clinical outcomes or cost-effective, guideline-concordant care has not been determined [9]. Currently, Ireland has no national LBP clinical pathway, although development has commenced.

Respondents’ expressed concern about the Irish healthcare system’s capacity for change; the underlying reasons warrant exploration, as well as whether CS physiotherapists, experts in MSK management, feel enabled to influence policy, funding and health system changes to improve MSK healthcare [29, 30]. Does the under-representation of HSCPs in health leadership roles [16] constrain their ability to effect meaningful reform and contribute to the persistence of an ineffective model of LBP care unsupported by best-practice evidence? Respondents considered their inability to access timely clinical imaging and consultant opinion in PC settings as leading to inefficiency and delayed care. Clinical specialist physiotherapists in Ireland remain reliant on medical teams for clinical imaging requests as ionising radiation prescription remains outside their scope of practice, despite the advocacy of the professional representative body, perhaps reflective of a lack of HSCP influence at health policy level.

Respondents envisaged that community MSK triage clinics will enhance collaborative working with PC physiotherapists, but identify barriers to be addressed to optimise successful implementation, including physical and IT infrastructure challenges and alienation from consultant teams. The Irish healthcare system has specific challenges to developing integrated PC teams owing to a complex public-private workforce mix. Just 40% of the population are entitled to free care from GPs, largely self-employed independent contractors, commonly in private premises independent of the wider PC MDT [31]. This PC model in Ireland poses challenges for the appointment of physiotherapists as first contact practitioners (FCPs) to reduce the MSK burden on GP services in the face of growing concern regarding GP shortages [31]. In the UK, FCP roles facilitate early access for patients to high-quality MSK advice in PC, with reduced onward referral to orthopaedics and low requirement for imaging [32]. Respondents’ concerns regarding reduced learning opportunities in community locations could be addressed through enhanced formal training and support; their reports that training is largely self-directed, self-funded and on their own time echoes the known deficit of standardised training for physiotherapists undertaking specialist roles [33, 34]. Unlike nursing colleagues, Irish physiotherapists, undertaking what are essentially advanced practice roles, do not have formal recognition of this responsibility. Development of advanced practice roles, with a clear educational pathway and independent prescribing of ionising radiation, has the potential to enable efficiencies in management of MSK disorders in PC.

Respondents’ recognition of the complex multidimensional nature of LBP-associated disability and of social and policy factors as determinants of such disability is reflected by the theme ‘need for a multisectoral approach to spine health’. The socioeconomic burden of LBP-associated disability and work-absence could be ameliorated by a collaborative multisectoral approach across health, social care, occupational and legal sectors [3], but this complex area warrants exploration of the issues, solutions and implementation challenges in the Irish context. Less challenging is respondents’ call for enhanced community exercise initiatives, which could form the foundation of a proactive public health approach to MSK health, focusing on health promotion and disease prevention, similar to that being implemented for other non-communicable diseases [35, 36].

## Strengths and limitations

Although previous research has sought physiotherapists’ views on various clinical aspects of LBP care (e.g. clinical guidelines [37], clinical reasoning [38] and specific treatment approaches [39]), this is the first study to document their opinions on broader health system influences on LBP care. Our findings are timely in informing the implementation of community-based MSK interface services and the new national LBP clinical pathway, currently in development. This study benefitted from being a national survey with a higher-than-average online survey response rate leading to qualitative data from 34 clinicians [40, 41], SRQR compliance, and the use of independent coding for rigour. Qualitative analysis of open-ended survey questions is limited by inability to explore meanings with participants; findings that would benefit from further examination include CS physiotherapists’ impression of health service resistance to change and their role as change agents in achieving improved organisational/ health system MSK healthcare. Nevertheless, the breadth of ideas presented attests to the depth of information provided.

## Conclusion

Health systems globally are struggling to manage the growing socio-economic and patient burden of LBP. In this study CS physiotherapists identify deficiencies in Irish LBP health services, including the persistence of a biomedical model, over-reliant on SC services. They advocate for investment in comprehensive PC MDTs to enable best practice, enhanced integration across PC and SC services, the development of a national clinical pathway, and a multisectoral approach to address the complexity of LBP-associated disability. Respondents identify concerns to be addressed to enable successful implementation of community-based MSK triage clinics, including infrastructure challenges, alienation from hospital consultant teams and access to clinical imaging. Specialist physiotherapists have valuable insights into the design and development of MSK healthcare quality improvement, which through meaningful clinical engagement may be used to effect positive change at clinician, organisation and systems levels to ensure best treatment and care for patients.

## Ethical approval

Ethics approval was granted by the UCD Human Research Ethics Committee (LS-E-22-108)

## Data Availability

All data produced in the present study are available upon request to the authors.

## Acknowledgements

The authors thank all respondents for taking the time to complete the survey and the National Musculoskeletal Physiotherapy Triage Initiative for their kind assistance with survey distribution.

## References

[1] Hurwitz EL, Randhawa K, Yu H, Cote P, Haldeman S. The Global Spine Care Initiative: a summary of the global burden of low back and neck pain studies. Eur Spine J. 2018; 27(Suppl 6):796–801. 10.1007/s00586-017-5432-9

[2] Wu A, March L, Zheng X, Huang J, Wang X, Zhao J, et al. Global low back pain prevalence and years lived with disability from 1990 to 2017: estimates from the Global Burden of Disease Study 2017. Ann Transl Med. 2020;8(6):299. 10.21037/atm.2020.02.175

[3] Foster NE, Anema JR, Cherkin D, Chou R, Cohen SP, Gross DP, et al. Prevention and treatment of low back pain: evidence, challenges, and promising directions. Lancet. 2018;391(10137):2368–83. 10.1016/S0140-6736(18)30489-6

[4] Liu S, Wang B, Fan S, Wang Y, Zhan Y, Ye D. Global burden of musculoskeletal disorders and attributable factors in 204 countries and territories: a secondary analysis of the Global Burden of Disease 2019 study. BMJ Open. 2022;12(6):e062183. 10.1136/bmjopen-2022-062183

[5] Morris R, Smith M. Demand for medical consultants and specialists to 2028 and the training pipeline to meeet demand: a high level stakeholder informed analysis. HSE-National Doctors Training and Planning 2020. Available from: https://www.hse.ie/eng/staff/leadership-education-development/met/plan/demand-for-medical-consultants-and-specialists-to-2028-november-updates-v2.pdf Accessed 1st June, 2023.

[6] Madanat R, Makinen TJ, Ryan D, Huri G, Paschos N, Vide J, et al. The current state of orthopaedic residency in 18 European countries. Int Orthop. 2017;41(4):681–7. 10.1007/s00264-017-3427-0

[7] Model of care for Rheumatology in Ireland. HSE, National Clinical Programme for Rheumatology; 2018. Available from: https://www.hse.ie/eng/about/who/cspd/ncps/rheumatology/achievements/model-of-care-for-rheumatology-in-ireland.pdf Accessed 1st June, 2023.

[8] Fennelly O, Blake C, FitzGerald O, Breen R, Ashton J, Brennan A, et al. Advanced practice physiotherapy-led triage in Irish orthopaedic and rheumatology services: national data audit. BMC Musculoskelet Disord. 2018;19(1):181. 10.1186/s12891-018-2106-7

[9] Murphy C, French H, McCarthy G, Cunningham C. Clinical pathways for the management of low back pain from primary to specialised care: a systematic review. Eur Spine J. 2022;31(7):1846–65. 10.1007/s00586-022-07180-4

[10] Traeger AC, Buchbinder R, Elshaug AG, Croft PR, Maher CG. Care for low back pain: can health systems deliver? Bull World Health Organ. 2019;97(6):423–33. 10.2471/BLT.18.226050

[11] Community Healthcare Organisations-Report and Recommendations of the Integrated Service Area Review Group. HSE Ireland; 2014. Available from: https://www.hse.ie/eng/services/publications/corporate/choreport.html Accessed 8th January, 2023

[12] Darker C. Integrated Healthcare in Ireland- A Critical Analysis and a Way Forward. The Adelaide Health Foundation, Ireland; 2013. Availanle from: https://www.tcd.ie/medicine/public_health_primary_care/assets/pdf/Integrated-Care-Policy-LR.pdf Accessed 12th May, 2023

[13] Jorm C, Hudson R, Wallace E. Turning attention to clinician engagement in Victoria. Aust Health Rev. 2019(43):123–5. 10.1071/AH17100

[14] Nilsen P, Seing I, Ericsson C, Birken SA, Schildmeijer K. Characteristics of successful changes in health care organizations: an interview study with physicians, registered nurses and assistant nurses. BMC Health Serv Res. 2020;20(1):147. 10.1186/s12913-020-4999-8

[15] Shaikh U, Lachman P, Padovani AJ, McCarthy SE. The care and keeping of clinicains in quality improvement. Int J Qual Health Care. 2020;32(7):480–5. 10.1093/intqhc/mzaa071

[16] Hanafin S, Shannon M, Jayachandran MS, Reed J. Impacts of Health and Social Care Professionals Leadership in Publicly-Funded Health Services in Ireland. Journal of Social Science & Allied Health Professions. 2020. Available from: http://hdl.handle.net/10147/628182 Accessed 25th May, 2023.

[17] O’Brien BC, Harris IB, Beckman TJ, Reed DA, Cook DA. Standards for reporting qualitative research: a synthesis of recommendations. Acad Med. 2014;89(9):1245–51. 10.1097/ACM.0000000000000388

[18] Gale NK, Heath G, Cameron E, Rashid S, Redwood S. Using the framework method for the analysis of qualitative data in multi-disciplinary health research. BMC Med Res Methodol. 2013;13(117). 10.1186/1471-2288-13-117

[19] Braun V, Clarke V. Successful qualitative research: a practical guide for beginners. London: SAGE Publications Ltd.; 2013. Chapter 2, Ten fundamentals of qualitative research; p. 19-41.

[20] Childs JD, Fritz J, Wu SS, Flynn TW, Wainner RS, Robertson EK, et al. Implications of early and guideline adherent physical therapy for low back pain on utilization and costs. BMC Health Serv Res. 2015; 15(150). 10.1186/s12913-015-0830-3

[21] Fritz JM, Joshua A, Brennan G. Does Adherence to the Guideline Recommendation for Active Treatments Improve the Quality of Care for Patients With Acute Low Back Pain Delivered by Physical Therapists? Medical Care. 2007;45(10):973–80. 10.1097/MLR.0b013e318070c6cd

[22] Fifer SK, Choundry NK, Brod M, Hsu E, Milstein A. Improving adherence to guidelines for spine pain care: what tools could support primary care clinicians in conforming to guidelines? BMJ Open Qual. 2022;11(3). 10.1136/bmjoq-2022-001868

[23] Slade SC, Kent P, Patel S, Bucknall T, Buchbinder R. Barriers to Primary Care Clinician Adherence to Clinical Guidelines for the Management of Low Back Pain: A Systematic Review and Metasynthesis of Qualitative Studies. Clin J Pain. 2016;32(9):800–16. 10.1097/AJP.0000000000000324

[24] Eighan J, Walsh B, Smith S, Wren MA, Barron S, Morgenroth E. A profile of physiotherapy supply in Ireland. Ir J Med Sci. 2019;188(1):19–27. 10.1007/s11845-018-1806-1

[25] Dutmer AL, Schiphorst Preuper HR, Soer R, Brouwer S, Bultmann U, Dijkstra PU, et al. Personal and Societal Impact of Low Back Pain: The Groningen Spine Cohort. Spine (Phila Pa 1976). 2019;44(24):E1443-E51. 10.1097/BRS.0000000000003174

[26] Briggs AM, Persaud JG, Deverell ML, Bunzli S, Tampin B, Sumi Y, et al. Integrated prevention and management of non-communicable diseases, including musculoskeletal health: a systematic policy analysis among OECD countries. BMJ Glob Health. 2019;4(5):e001806. 10.1136/bmjgh-2019-001806

[27] Huckel Schneider C, Parambath S, Young JJ, Jain S, Slater H, Sharma S, et al. From Local Action to Global Policy: A Comparative Policy Content Analysis of National Policies to Address Musculoskeletal Health to Inform Global Policy Development. Int J Health Policy Manag. 2023;12(Issue1): 1–20. 10.34172/ijhpm.2022.7031

[28] Johnson CD, Haldeman S, Chou R, Nordin M, Green BN, Cote P, et al. The Global Spine Care Initiative: model of care and implementation. Eur Spine J. 2018;27(Suppl 6):925–45. 10.1007/s00586-018-5720-z

[29] Finucane LM, Stokes E, Briggs AM. Its everyone’s responsibility: Responding to the global burden of musculoskeletal health impairment. Musculoskelet Sci Pract. 2023;64:102743. 10.1016/j.msksp.2023.102743

[30] Lewis JS, Stokes EK, Gojanovic B, Gellatly P, Mbada C, Sharma S, et al. Reframing how we care for people with persistent non-traumatic musculoskeletal pain. Suggestions for the rehabilitation community. Physiotherapy. 2021;112:143–9. 10.1016/j.physio.2021.04.002

[31] Connolly S, Brick A, O’Neill C, O’Callaghan M. An analysis of the Primary care systems of Ireland and Northern Ireland. ESRI Research Series 137, Dublin: ERSI, 10.26504/rs137

[32] Downie F, McRitchie C, Monteith W, Turner H. Physiotherapist as an alternative to a GP for musculoskeletal conditions: a 2-year service evaluation of UK primary care data. Br J Gen Pract. 2019;69(682):e314–e20. 10.3399/bjgp19X702245

[33] Fennelly O, Desmeules F, O’Sullivan C, Heneghan NR, Cunningham C. Advanced musculoskeletal physiotherapy practice: Informing education curricula. Musculoskelet Sci Pract. 2020;48:102174. 10.1016/j.msksp.2020.102174

[34] Skinner EH, Haines KJ, Hayes K, Seller D, Toohey JC, Reeve JC, et al. Future of specialised roles in allied health practice: who is responsible? Aust Health Rev. 2015;39:255–9. 10.1071/AH14213

[35] HSE National Chronic Disease Management Programme [Available from: https://www.hse.ie/eng/about/who/gmscontracts/2019agreement/chronic-disease-management-programme/ Accessed 2nd July, 2023

[36] Budreviciute A, Damiati S, Sabir DK, Onder K, Schuller-Goetzburg P, Plakys G, et al. Management and Prevention Strategies for Non-communicable Diseases (NCDs) and Their Risk Factors. Front Public Health. 2020;8:574111. 10.3389/fpubh.2020.574111

[37] van den Heuvel C, van der Horst J, Winkelhorst E, Roelofsen E, Hutting N. Experiences, barriers and needs of physiotherapists with regard to providing self-management support to people with low back pain: A qualitative study. Musculoskelet Sci Pract. 2021;56:102462. 10.1016/j.msksp.2021.102462

[38] Widerstrom B, Rasmussen-Barr E, Bostrom C. Aspects influencing clinical reasoning and decision-making when matching treatment to patients with low back pain in primary healthcare. Musculoskelet Sci Pract. 2019;41:6–14. 10.1016/j.msksp.2019.02.003

[39] Ayre J, Jenkins H, McCaffery KJ, Maher CG, Hancock MJ. Physiotherapists have some hesitations and unmet needs regarding delivery of exercise programs for low back pain prevention in adults: A qualitative interview study. Musculoskelet Sci Pract. 2022;62:102630. 10.1016/j.msksp.2022.102630

[40] Cook C, Heath F, Thompson RL. A Meta-Analysis of Response Rates in Web- or Internet-Based Surveys. Educ Psychol Meas. 2000;60(6):821–36. 10.1177/00131640021970934

[41] Meyer VM, Benjamens S, Moumni ME, Lange JFM, Pol RA. Global Overview of Response Rates in Patient and Health Care Professional Surveys in Surgery: A Systematic Review. Ann Surg. 2022;275(1):e75–e81. 10.1097/SLA.0000000000004078

